# GWAS and 3D chromatin mapping identifies multicancer risk genes associated with hormone-dependent cancers

**DOI:** 10.1101/2024.07.14.24310389

**Authors:** Isela Sarahi Rivera, Juliet D. French, Mainá Bitar, Haran Sivakumaran, Sneha Nair, Susanne Kaufmann, Kristine M. Hillman, Mahdi Moradi Marjaneh, Jonathan Beesley, Stacey L. Edwards

**Author notes:** These authors jointly supervised the work (Juliet D. French, Jonathan Beesley, Stacey L. Edwards).

## Abstract

Hormone-dependent cancers (HDCs) share several risk factors, suggesting a common aetiology. Using data from genome-wide association studies, we showed spatial clustering of risk variants across four HDCs (breast, endometrial, ovarian and prostate cancers), contrasting with genetically uncorrelated traits. We identified 44 multi-HDC risk regions across the genome, defined as overlapping risk regions for at least two HDCs: two regions contained risk variants for all four HDCs, 13 for three HDCs and 28 for two HDCs. Integrating GWAS data, epigenomic profiling and high-resolution promoter capture HiC maps from diverse cell line models, we annotated 53 candidate risk genes at 22 multi-HDC risk regions. These targets were enriched for established genes from the COSMIC Cancer Gene Census, but many had no previously reported pleiotropic roles. Additionally, we pinpointed lncRNAs as potential HDC targets and identified risk alleles in several regions that altered transcription factors motifs, suggesting regulatory mechanisms. Known drug targets were over-represented among the candidate multi-HDC risk genes, implying that some may serve as targets for therapeutic development or facilitate the repurposing of existing treatments for HDC. Our comprehensive approach provides a framework for identifying common target genes driving complex traits and enhances understanding of HDC susceptibility.

**AUTHOR SUMMARY:** While hormone-dependent cancers (HDCs) share several risk factors, our understanding of the complex genetic interactions contributing to their development is limited. In this study, we leveraged large-scale genetic studies of cancer risk, high-throughput sequencing methods and computational analyses to identify genes associated with four HDCs: breast, endometrial, ovarian and prostate cancers. We identified known cancer genes and discovered many that were not previously linked to cancer. These findings are significant because identifying genes associated with risk of multiple cancer types can enhance the gene mapping accuracy and highlight new therapeutic targets.

## INTRODUCTION

Breast, endometrial, ovarian and prostate cancers are hormone-dependent cancers (HDCs) that together account for up to 30% of new cancer diagnoses each year (1). These cancers share several environmental, behavioural and genetic risk factors, suggesting a common aetiology (2, 3). This premise is supported by genome-wide association studies (GWAS) which have identified hundreds of cancer-specific risk loci (4–7) and multiple pleiotropic loci associated with at least two HDCs (8–11). The detection of pleiotropic loci suggests that shared genetic factors likely contribute to polygenic risk of HDCs and raises the possibility of common driver genes and biological pathways.

A key aim of post-GWAS is to identify the target gene(s) that are affected at each GWAS region. This is complicated by the fact that most risk variants reside in noncoding regions of the genome making it difficult to interpret how they contribute to cancer susceptibility (12). Target gene mapping for pleiotropic loci typically relies on statistical approaches such as expression quantitative trait loci (eQTL) and transcriptome-wide association studies (TWAS) (9, 10, 13). However, these methods are limited by small sample sizes which reduces power, the use of steady-state gene expression and the lack of data from relevant tissues. Orthologous functional assays provide complementary mapping approaches to better define regulatory variants and connect them to their target genes.

## RESULTS AND DISCUSSION

While most GWAS variants are dispersed across the genome, several genomic regions harbour variants for multiple cancer types, suggesting a common mechanism underlying susceptibility. Independent signals for HDC were obtained from large-scale GWAS, meta-analyses and fine-mapping (196 breast (4), 17 endometrial (5), 60 ovarian (6) and 258 prostate cancer variants (7)). To assess whether HDC variants were more frequently positioned together than would be expected by chance, we defined “clusters” when variants associated with two or more HDCs were co-localized within the same 100 kb window. We compared observed cluster frequency to a null distribution, generated by randomly shuffling variants one million times (see Methods), and recounted cluster occurrence in each random set. This resulted in a statistically significant increase in observed frequency (**Fig 1A**; *P* < 10^-6^, permutation test). As an additional comparison, index variants for four genetically uncorrelated traits (14) were obtained from the Open Target Genetics portal (179 for coronary artery disease, 22 lung cancer, 38 Alzheimer’s disease and 94 Parkinson’s disease (15)). These traits showed similar genomic distributions to randomly generated background regions (**Fig 1A**), which provided support that HDC risk regions co-localize in the genome and that these regions likely contain pleiotropic variants and common target genes.

To identify candidate multi-HDC risk (mHDCR) regions, variants for each HDC were extended by 0.5 Mb on both sides, resulting in cancer-specific risk regions. mHDCR regions were then defined as overlapping cancer-specific risk regions for at least two HDCs (**Fig 1B**). In total, 44 mHDCR regions were identified across the genome (**Fig 1C** and **Table S1**). Two mHDCR regions contained risk variants for all four HDCs, 13 mHDCR regions for three HDCs (seven breast-ovarian-prostate, four breast-endometrial-prostate and two endometrial-ovarian-prostate) and 28 mHDCR regions for two cancer types (twenty breast-prostate, four breast-ovarian, two ovarian-prostate, one endometrial-ovarian and one endometrial-prostate) (**Fig 1D**). Due to much larger sample sizes and fine-mapping analyses (4, 7) most breast and prostate mHDCR regions contain at least three independent signals per cancer type (**Fig 1E**).

**Fig 1.**
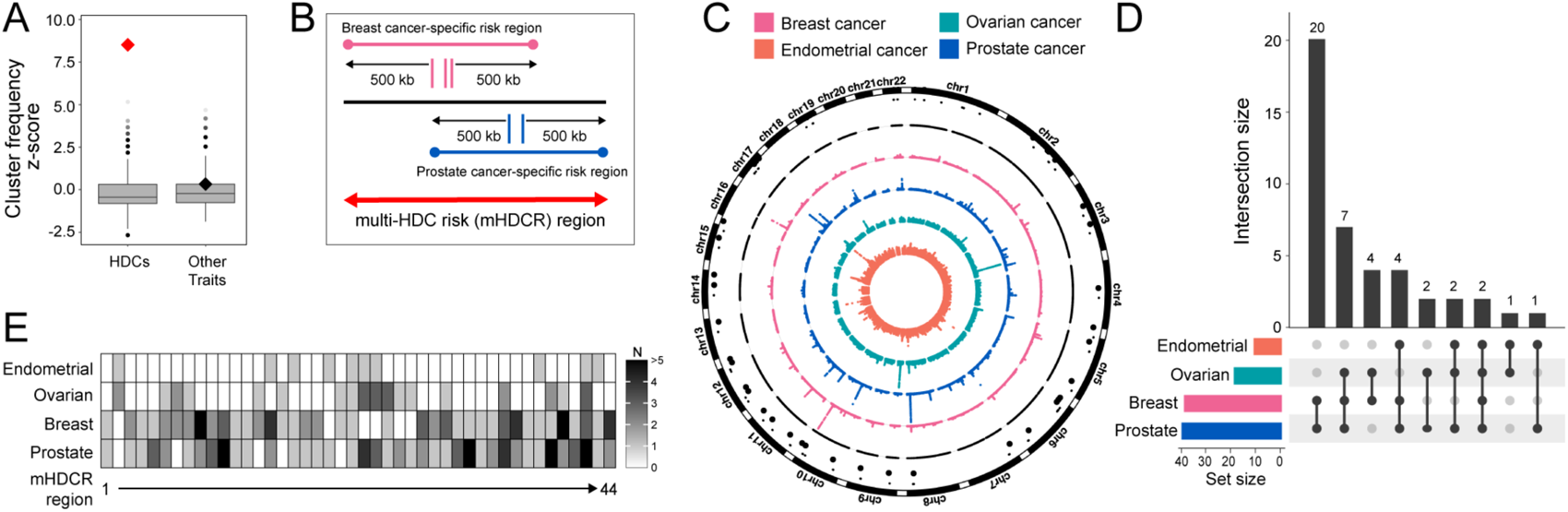
GWAS variants from four HDCs cluster in mHDCR regions. **(A).** Boxplots show the null distribution of z-scores from 10^6^ random permutations of the positions of variants associated with four HDCs (breast, endometrial, ovarian and prostate cancers), and four genetically uncorrelated traits (coronary artery disease, lung cancer, Alzheimer’s disease and Parkinson’s disease). The true cluster count score is indicated with a diamond, boxes represent the median and interquartile range and whiskers represent upper and lower quartiles of the null distribution. **(B).** Schematic of a hypothetical mHDCR region (red arrow) containing variants for breast cancer (pink) and prostate cancer (blue) which were extended to generate the cancer-specific risk regions. **(C).** Circus plot providing an overview of the association results. Dots in the four coloured rings correspond to genome-wide significant (-log_10_ (5 x 10^-8^)) GWAS variants identified for breast, endometrial, ovarian and prostate cancers, ordered by chromosomal position. The black ring denotes chromosomes (chr) 1-22 and the black dots specify the 44 mHDCR regions. **(D).** Upset plot to illustrate the number of mHDCR regions shared between the four HDCs. The vertical barplot represents the total number of mHDCR regions in each HDC combination. Points and lines in the matrix visualize these connections, and the colored horizontal bars are the total number of mHDCRs in each HDC. **(E).** Heatmap showing the number (N) of GWAS variants in each mHDCR region per cancer type.

Candidate mHDCR gene(s) within the 44 mHDCR regions were identified using a multistep computational approach (**Fig 2A**). For endometrial, ovarian and prostate cancers, each variant was expanded to include all candidate variants (r^2^ ≥ 0.8; 1000 Genomes phase 3 version 5 reference) (**Table S2**). For breast cancer, the candidate causal variants from the recent fine-mapping study were used (4). We annotated each mHDCR region with protein-coding genes from GENCODE (v37 basic) and intersected all candidate variants with genomic annotations (exons, promoters defined as transcription start site (TSS) ± 2 kb and intronic and intergenic regions; **Fig S1A**). Exonic variants were filtered using Ensembl variant effect predictor (VEP) (16) which identified one frameshift and 33 moderate-impact missense and untranslated region (UTR) variants (**Fig S1B** and **Table S3**). Seven potential splicing variants were also detected with SpliceAI (17) and MaxEntScan (18). The promoter and intronic variants were further explored for regulatory functions using RegulomeDB (19). Sixty-seven variants had strong regulatory potential (score > 0.5, **Fig S1C** and **Table S4**), suggesting these variants are more likely to be functional. Previous studies have performed functional assays for some of the noncoding variants in individual HDCs (20–23). For example, Lawrenson et al used 3D mapping and reporter assays to show that breast and ovarian cancer risk variants at 19p13 influence distal enhancers, which in turn regulate *ABHD8* expression (21). Furthermore, Stegeman et al used reporter assays to show a prostate cancer risk variant alters microRNA binding to the *MDM4* 3’UTR (22). These studies provide additional support that the variants can impact target gene expression through various mechanisms.

A first-pass analysis identified 17 candidate mHDCR genes (at 11 mHDCR regions) that have exonic, promoter or potential splicing variants associated with two or more HDCs (**Fig 2B, Fig S1D** and **Table S5**). Of these, six are cancer genes based on information from the Catalogue of Somatic Mutations in Cancer (COSMIC (24)), the remaining genes are potentially new pleiotropic HDC risk genes. One example is the *PAX9* transcription factor (TF; mHDCR region 18) (25) which has a missense breast cancer risk variant plus promoter and intronic breast and prostate cancer risk variants (**Fig 2C**). Rs2236007 (breast) and rs12882923 (prostate) were the top-ranked variants on RegulomeDB and mapped to transcriptionally active open chromatin (defined by H3K4me3 marks and DNase-seq) in breast and prostate cells. Previous studies in breast cells showed that the rs2236007 risk *g*-allele reduced *PAX9* promoter activity (26), via recruitment of the suppressive TF EGR1 and increased breast cancer cell growth (27). Furthermore, rs12882923 maps to a POL2RA binding site and the DNA motif of multiple TFs in prostate epithelium (**Fig 2C**), suggesting the variant has functional impact.

**Fig 2.**
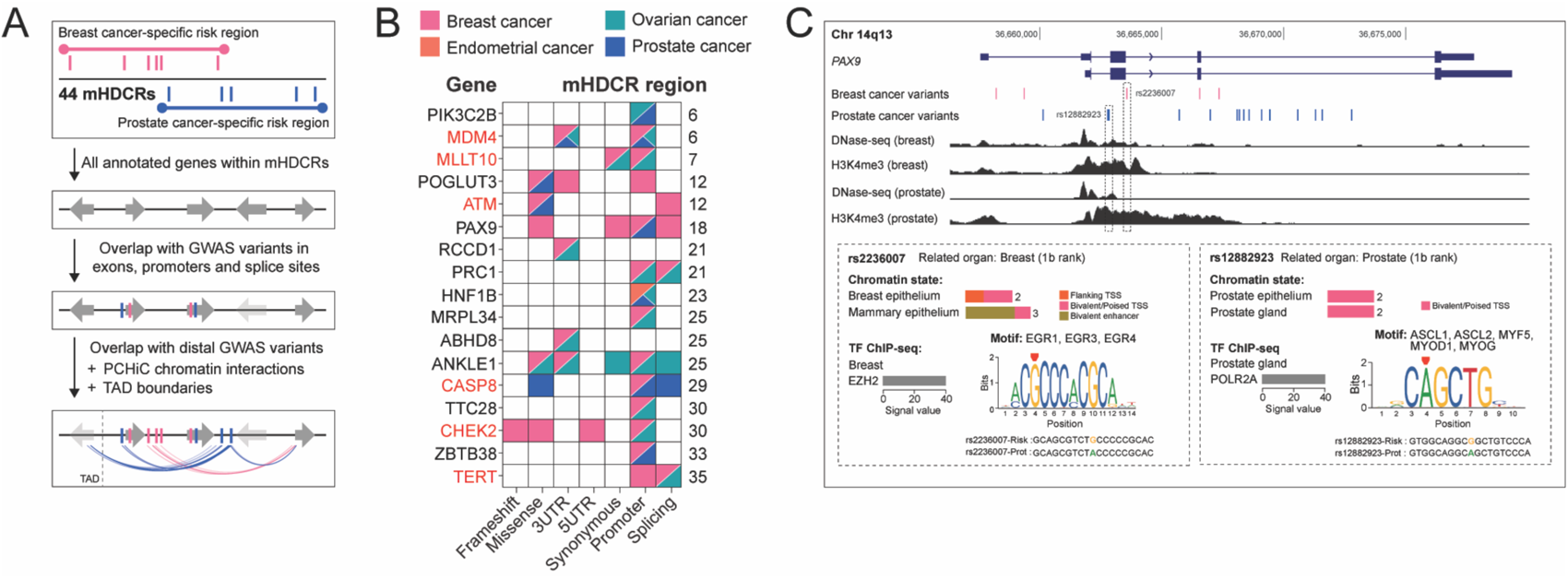
Identification of candidate mHDCR genes. **(A).** Schematic of the stepwise computational pipeline used to identify candidate mHDCR genes. **(B).** Summary of candidate mHDCR genes after overlap with variants in gene exons, promoters and splice sites that are associated with two or more HDCs (pink-breast cancer; orange-endometrial cancer; teal-ovarian cancer; blue-prostate cancer). The left y-axis shows the annotated gene names, the right y-axis denotes the mHDCR region. The red text highlights known cancer genes (from COSMIC) at the mHDCR regions. The x-axis provides the location and/or functional annotation of the HDC risk variants. Frameshift, missense, synonymous-variants located in gene exons; 3UTR/5UTR-variants; Promoter-variants located in gene promoters defined as TSS ± 2 kb; Splicing-variants in gene introns that are predicted to alter splicing. **(C).** WashU genome browser (hg38) showing GENCODE annotated genes (blue). The risk variants are shown as pink (breast cancer) and blue (prostate cancer) vertical lines. The DNase-seq and H3K4me3 tracks from breast and prostate cells are shown as black histograms. The dashed gray outlines highlight the functional variants. Insets: RegulomeDB v2.2 analysis for rs2236007 and rs12882923 including heuristic ranking scores, presence in promoters or enhancers (chromatin state) and sequences affecting the binding of TFs (TF ChIP-seq) and DNA motifs.

Most HDC risk variants are noncoding (**Fig S1A**), and a subset will map to DNA regulatory elements that modulate gene transcription through long-range chromatin interactions (28). To compile a comprehensive list of candidate mHDCR genes, we next intersected the distal intronic and intergenic HDC variants with the 44 mHDCR regions. High-resolution promoter capture HiC (PCHiC) was used to assign distal regulatory variants to their target gene promoters (**Fig 3A**). PCHiC probes were designed to 2774 HindIII fragments containing 3096 GENCODE gene promoters that fall within the 44 mHDCR regions (**Table S6**). PCHiC data was derived from twelve breast, endometrial, ovarian and prostate non-tumorigenic and cancer cell lines, as described previously (29) (**Fig S2A** and **Table S7**). Using an interaction score threshold (CHiCAGO score ≥ 5) we detected 8-19,000 high-confidence interactions per cell type across the 44 mHDCR regions with strong correlation between replicates (**Fig 3B** and **Fig S2B**). We prioritised the PCHiC interactions based on topologically associating domain (TAD) boundaries (30). To show that a published set of TAD annotations marked regions of increased regulatory activity, we examined the relationship between our PCHiC interactions and TAD boundaries. We observed a significant enrichment of PCHiC interactions within TAD boundaries, compared to randomly placed TAD boundaries (*P* = 0.0297, permutation test), suggesting that the published TAD boundaries mark the limits of most PCHiC interactions in our HDC cell lines (**Fig 3C**).

**Fig 3.**
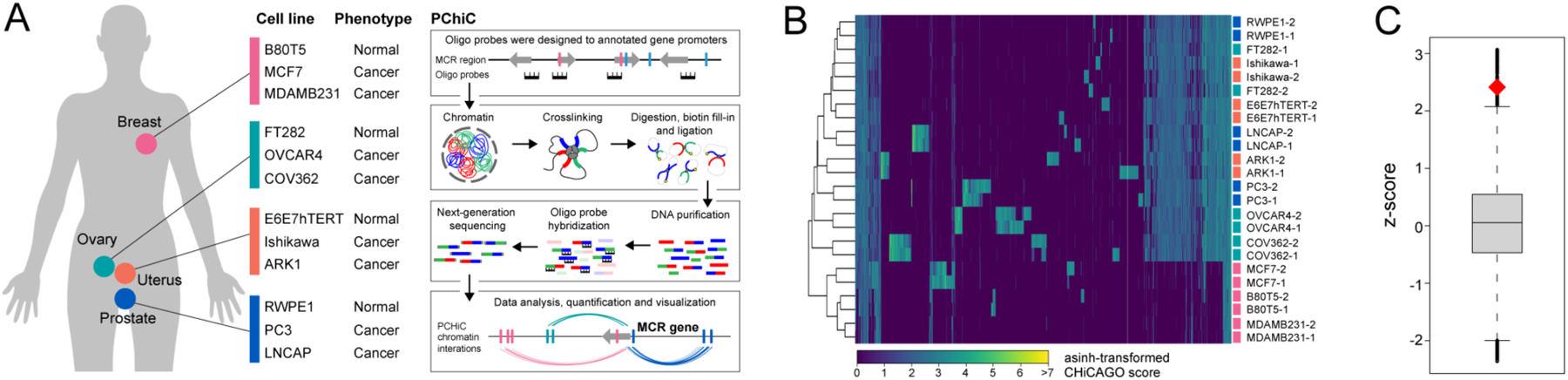
PCHiC in HDC cell lines. **(A).** Schematic of PCHiC experimental approach. **(B).** Agglomerative hierarchical clustering for the PCHiC in the twelve cell lines. **(C).** Enrichment of PCHiC interactions within TADs. The boxplot shows the null distribution z-scores generated from randomly permuting the TAD boundary positions within mHDCR regions (n = 10^5^) and counting the number of interactions which fall within randomized TAD boundaries. The diamond indicates the z-score of the count of interactions which fall within observed TAD boundaries. The box represents the median and interquartile range, whiskers represent upper and lower quartiles of the null distribution.

The combination of variant intersection, PCHiC and TAD boundaries identified 53 candidate mHDCR genes (at 22 mHDCR regions) associated with two or more HDCs (**Fig 4A** and **Table S5**). Ten candidate mHDCR genes were associated with three cancer types (seven breast-ovarian-prostate and three endometrial-ovarian-prostate) and 43 candidate mHDCR genes with two cancer types (seventeen breast-ovarian, fifteen breast-prostate, eight endometrial-prostate and three ovarian-prostate) (**Fig 4B**). The expression of all candidate mHDCR genes in the relevant tissue or cancer type was estimated using GTEx and TCGA RNA-seq data (**Fig S3**). Twelve are cancer genes from COSMIC, doubling the number of candidate HDC cancer genes when regulatory variants are taken into account (**Fig 4A**). As one example, *TCF7L2* (transcription factor 7 like 2) has 5UTR, promoter and intronic prostate cancer variants that are associated with the risk of developing aggressive prostate cancer (31) (**Fig 4C**). However, our combined analysis showed that *TCF7L2* also has intronic breast and ovarian cancer risk variants plus distal breast cancer risk variants which loop to and may regulate an alternate *TCF7L2* isoform (**Fig 4C**).

Notably, two-thirds of the genes in the final list required the inclusion of distal variants and PCHiC data to classify them as candidate mHDCR genes. Some are known pleiotropic cancer genes (e.g *MYC*, *CCND1;* **Fig S4**) supporting the validity of our approach, but many have only been statistically and/or functionally associated with individual HDCs and may represent new mHDCR genes. One example is *NWD1* (NACHT and WD repeat domain containing 1) at 19p13 (**Fig 4D**). Previous studies have detected variants at this region associated with breast and ovarian cancer risk and identified *ABHD8* and *ANKLE* as the likely target genes (21). Here we show that *NWD1*, located ∼580 kb from *ABHD8*, has distal ovarian and prostate cancer variants, some of which fall in regulatory elements that loop to and may regulate the *NWD1* promoter (**Fig 4D**). Overexpression of NWD1 is reported to promote prostate tumor progression by modulation of androgen receptor (AR) signaling (32). There is limited information about NWD1 function in ovarian cancer, but evidence suggests AR signaling also contributes to initiation and progression of this disease (33), providing a potential mechanism.

It is established that drug targets with genetic support (such as well-powered GWAS) are twice as likely to lead to approved drugs (34). To determine whether the candidate mHDCR genes were enriched for known drug targets, we mined the Citeline Pharmaprojects database (35). We found twelve candidate mHDCR genes that encode known preclinical, clinical phase and approved drug targets for various diseases (**Fig 4E** and **Table S8**). The overlap between known drug targets and the candidate mHDCR genes was statistically significant (**Fig 4F**; OR = 2.2, *P* = 0.028, Fisher’s Exact Test), indicating the value of using our pipeline to identify candidates for drug repositioning.

**Fig 4.**
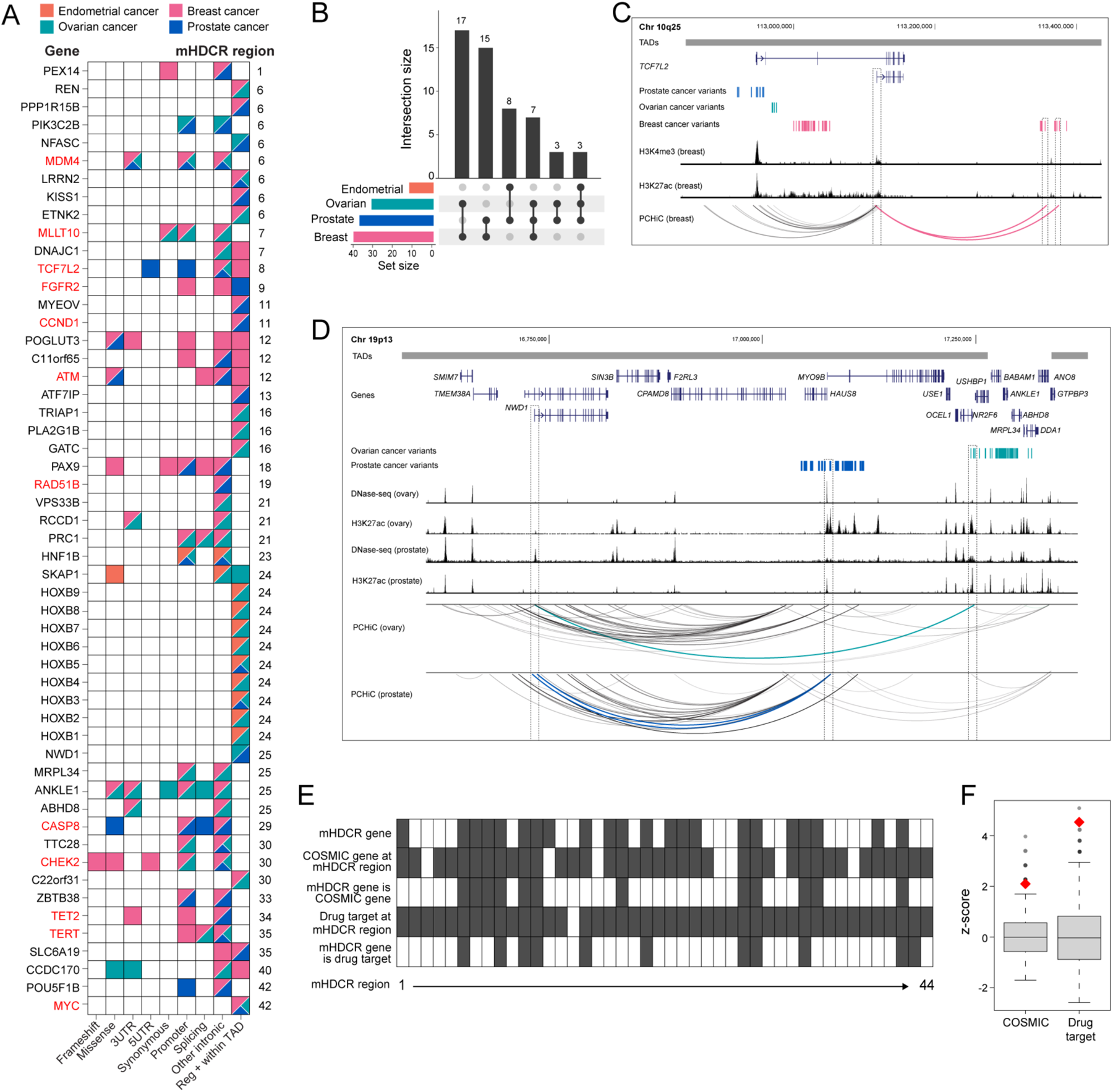
Annotation of candidate mHDCR genes. **(A).** Summary of all candidate mHDCR genes that are associated with two or more HDCs (pink-breast cancer; orange-endometrial cancer; teal-ovarian cancer; blue-prostate cancer). The left y-axis shows the annotated gene names, the right y-axis denotes the mHDCR region. The red text highlights known cancer genes (from COSMIC) at the mHDCR regions. The x-axis provides the location and/or functional annotation of the HDC risk variants. Frameshift, missense, synonymous-variants located in gene exons; 3UTR/5UTR-variants; Promoter-variants located in gene promoters defined as TSS ± 2 kb; Splicing-variants in gene introns that are predicted to alter splicing; Other intronic-variants located in gene introns and RegulomeDB scores indicate the variants are regulatory; Reg + within TAD-variants located outside any genes, RegulomeDB scores indicate the variants are regulatory, our PCHiC data show chromatin interactions between the variant and gene promoters and the interactions are within defined TAD boundaries. **(B).** Upset plot to illustrate the number of candidate mHDCR genes shared between the four HDCs. The vertical barplot represents the total number of candidate mHDCR genes in each HDC combination. Points and lines in the matrix visualize these connections, and the colored horizontal bars are the total number of candidate mHDCR genes in each HDC. **(C).** WashU genome browser (hg38) showing TADs as horizontal gray bars above GENCODE genes (blue). The risk variants are shown as blue (prostate cancer), teal (ovarian cancer) and pink (breast cancer) vertical lines. The H3K4me3 and H3K27ac tracks from breast cells are shown as black histograms. CHiCAGO-scored interactions are shown as colored arcs. The dashed gray outlines highlight distal breast cancer variants and the target gene (*TCF7L2*). **(D).** WashU genome browser (hg38) showing TADs as horizontal gray bars above GENCODE genes (blue). The risk variants are shown as teal (ovarian cancer) and blue (prostate cancer) vertical lines. The DNase-seq and H3K27ac tracks from ovary and prostate cells are shown as black histograms. CHiCAGO-scored interactions are shown as colored arcs. The dashed gray outlines highlight the likely functional variants and the target gene (*NWD1*). **(E).** Summary of identified candidate mHDCR genes at the 44 mHDCR regions for HDCs. Filled boxes represent presence of the feature according to the row title. **(F).** Enrichment of COSMIC genes and genes which encode known drug targets in candidate mHDCR genes. The boxplots represent the null distribution z-scores following 10^5^ random permutations of category labels (COSMIC or drug target) of genes at mHDCR regions, then recounting the number of predicted mHDCR genes belonging to the category. The z-scores for the true observed counts are shown as red diamonds. Boxes represent the median and interquartile range and whiskers represent upper and lower quartiles of the null distribution.

Our approach was unable to identify candidate mHDCR genes at 22 regions. There are several possible explanations for this, the simplest being that the genetic effects at these mHDCR regions are only associated with risk of a specific HDC. Furthermore, we focused on identifying protein-coding genes, but studies by ourselves (and others) show that long noncoding RNAs (lncRNAs) are also transcribed from cancer risk regions and can have important roles in tumorigenesis (36–40). Indeed, when we intersected the HDC risk variants with annotated lncRNAs at the 44 mHDCR regions, we identified 21 lncRNAs associated with two or more HDCs, including two cancer-related lncRNAs that were the only targets identified at mHDCR regions 27 and 32 (**Fig S5**) (41, 42). Given most cell-type specific lncRNAs are not annotated in current databases, we expect that additional novel lncRNAs will contribute to risk at other mHDCR regions. We acknowledge that some distal gene interactions may have been missed due to intrinsic biases in the PCHiC. For example, false negatives may result from the lack of suitable baits for selected promoters, the reduced resolution for short-range interactions or due to the transient and cell type-specific nature of chromatin interactions. It is also important to note that interactions between risk variants and gene promoters do not infer causality and that follow-up studies are required to interpret GWAS results.

In summary, our study expands the repertoire of risk regions and candidate risk genes for HDCs, providing further insights into the complex genetic architecture and biology underpinning HDC susceptibility. The candidate mHDCR gene list was enriched for known cancer genes and drug targets, which provides support that other, less-well-characterized genes at mHDCR regions may play important roles in HDC development. Our approach highlights the value of performing integrative analyses of genetics and functional genomics to enhance pleiotropic cancer gene identification. Combined with future research that investigates functional mechanisms, our results may serve to redirect efforts to more promising targets for new drugs or allow drug repurposing for HDC treatment.

## MATERIALS AND METHODS

### Genomic interval operations

Genomic interval analyses were performed with BEDTools version 2.29.0 (43). To assess the genomic distribution of genetic signals, the *cluster* sub-command was used to assign IDs to GWAS variants overlapping 100 kb windows. Co-located variants for two or more traits were designated as clusters. For comparison, positions of the HDC variants were shuffled while maintaining chromosomal distribution using BEDTools *shuffle* sub-command, and clusters counted for each permutation. To explore the relationship between PCHiC loops and TADs, boundary positions within multi-cancer regions were overlapped with loops using the BEDTools *intersect* sub-command. For comparison, we generated null distributions by shuffling TAD boundary positions within MCRs and repeating 10,000 times. TAD size was maintained by circularising the MCR, such that if a TAD boundary was randomly placed beyond the MCR limits, it was moved to the start of the MCR. The background count was performed by intersecting loops with each iteration of randomly permuted boundary sets. The significance of these tests were defined by the number of times the count from random permutations was greater or equal to the observed overlap, divided by total number of permutations. For presentation, the null distribution was standardised to produce z-scores and shown as box plots.

### Variant annotation

Candidate causal variant rsIDs were submitted to the Variant Effect Predictor (VEP) web interface. Coding variants annotated as frameshift, missense, 3UTR, 5UTR or synonymous were prioritized as potentially functional with an impact on the associated transcript. Intronic variants with the potential to alter splicing were identified using 1) SpliceAI Delta scores ≥ 0.2, where delta can be interpreted as the probability that the variant is splice altering; or 2) absolute MaxEntScan maximum entropy scores > 0. The regulatory effects of all variants were assessed using RegulomeDB. Variants with a probability score greater than the median (0.55) were considered as potential functional. Genomic coordinates for topologically associating domains (TADs) were obtained from a recent publication (44). Annotation of variants with respect to genomic intervals (exons, introns, TADs, and promoters) and to link with candidate distal target genes were performed with the GenomicRanges and GenomicInteractions BioConductor packages. Genes with known roles in cancer were obtained from the COSMIC Cancer Gene Census, version 99 (24). Information about drug targets and indications was obtained from (35).

### Cell lines and culture conditions

MCF7 cells were cultured in RPMI medium with 10% (vol/vol) fetal bovine serum (FBS, Gibco), 1% (vol/vol) antibiotic/antimycotic (a/a, Gibco), 1mM sodium pyruvate (Gibco), 0.02M Hepes (pH 7.0-7.4) and 10 µg/mL human recombinant insulin (Gibco). MDA-MB-231 cells were grown in RPMI medium with 10% (vol/vol) FBS, 1% (vol/vol) a/a, 1 mM sodium pyruvate, 0.02M Hepes (pH 7.0-7.4). B80T5, ARK1, LNCAP and OVCAR4 cells were grown in RPMI medium with 10% (vol/vol) FBS and 1% (vol/vol) a/a. E6E7hTERT, Ishikawa and COV362 cells were grown in DMEM medium with 10% (vol/vol) FBS and 1% (vol/vol) a/a. PC3 cells were grown in DMEM:F12 medium with 10% (vol/vol) FBS and 1% (vol/vol) a/a. RWPE1 cells were grown in Gibco Keratinocyte-SFM Combo (Cat# 17005042) with 1% (vol/vol) a/a. FT282 cells were grown in DMEM:F12 (no Hepes) medium with 5% USG (serum replacement supplement). Cell lines were maintained at 37°C and 5% CO_2_, routinely tested for *Mycoplasma* and profiled for short tandem repeats.

### Biotinylated RNA bait library design and HiC libraries capture

To generate target regions for PCHiC, mHDCR regions were extended by 1Mb on either side, so that all possible promoter-enhancer interactions involving regions containing HDC risk variants were captured. All HindIII fragments containing annotated promoters within each extended region were identified. HindIII fragments previously captured by Javierre *et al* (45) were labelled ‘group1’ and those not captured but overlapping Ensembl annotated promoters (46) were labelled ‘group2’. Capture probes were designed to both ends of group1 and group2 HindIII fragments (2783 fragments and 4023 probes, respectively).

### Promoter capture HiC (PCHiC) library preparation and sequencing

PCHiC was performed on nine immortalized HDC cell lines. For prostate: one normal cell line (RWPE1), one androgen-dependent (LNCAP) and one castration-resistant (PC3) cell line. For ovarian: one normal ovarian cancer precursor cell line (FT282) and two high-grade serous ovarian cancer cell lines (OVCAR4 and COV362). For endometrial: one normal endometrial cell line (E6E7hTERT), one ER+ (Ishikawa) and one type II EC line (ARK1). For breast: we remapped our published PCHiC data to mHDCR regions [29]. PCHiC libraries were prepared from 4-5 x 10^7^ cells per library (two biological replicates per cell line using in-nucleus ligation (47)). The HiC libraries were amplified using the SureSelectXT ILM Indexing pre-capture primers (Agilent Technologies) with 8 PCR amplification cycles. Each HiC library (750 ng) was hybridized and captured individually using the SureSelectXT Target Enrichment System reagents and protocol (Agilent Technologies). After library enrichment, a post-capture PCR amplification step was carried out using SureSelectXT ILM Indexing post-capture primers (Agilent Technologies) with 14–16 PCR amplification cycles. PCHiC libraries were multiplexed and sequenced on NovoSeq 6000 S4 (Novogene, Singapore).

### PCHiC analysis

PCHiC reads were analysed with *HiCUP* (version 0.5.9). Raw sequencing reads were aligned to the hg38 human reference genome with *Bowtie2* version 2.2.9 and filtered to remove experimental artefacts (e.g. re-ligated, circularized, unligated fragments or fragments separated by less than 20 kb). Chromatin interactions were identified using CHiCAGO following generation of genome indices, HindIII restriction fragment digest files, and the bait map. The CHiCAGO pipeline was run independently for each library for quality control, followed by merging of replicates for subsequent analyses. Interactions with CHiCAGO score ≥ 5 were treated as high confidence for target gene analysis and visualisation.

## Data Availability

All data produced in the present study are available upon reasonable request to the authors.

## ACKNOWLEDGEMENTS

This work was supported by a grant from the Cancer Council Queensland (1156712). I.S.R. was supported by a QIMR Berghofer Maureen Stevenson PhD Scholarship, a QIMR Berghofer top-up scholarship and a QUT HDR Tuition fee sponsorship. J.D.F. was supported by a philanthropic donation from Isabel and Roderic Allpass and an NHMRC Investigator Grant (2016826). S.L.E. was supported by a NHMRC Senior Research Fellow (1135932). The results published here are in part based upon the data generated by the TCGA Research Network. The Genotype-Tissue Expression (GTEx) Project was supported by the Common Fund of the Office of the Director of the National Institutes of Health, and by NCI, NHGRI, NHLBI, NIDA, NIMH, and NINDS. Funding for open access charge: QIMR Berghofer offers institutional grants.

